# Epigenetic Aging and Associated Factors in Japanese People with HIV: A Protocol for a Single-Center Cross-Sectional Study

**DOI:** 10.64898/2026.09.04.26362252

**Authors:** Yui Tomo, Seowoong Jung, Yuka Kudo-Nagata, Ryoko Sekiya, Kazuaki Fukushima, Ai Kawana-Tachikawa, Ryo Nakaki, Akifumi Imamura, Ayu Kasamatsu

## Abstract

**Background:** Advances in antiretroviral therapy (ART) have transformed human immunodeficiency virus (HIV) infection into a manageable chronic condition. However, people with HIV (PWH) face an increased burden of age-related non-AIDS-defining illnesses. Epigenetic age (EA) and epigenetic age acceleration (EAA), based on DNA methylation, have emerged as objective indicators of biological aging. Although studies in Western countries suggest that EAA levels are elevated in PWH and may decline after ART initiation, evidence in the Japanese population remains scarce. This study aims to characterize EA and EAA in Japanese PWH and to evaluate their associations with the duration of ART, cumulative exposure to specific drug classes, frailty, lifestyle factors, and immunological, metabolic, and inflammatory markers.

**Methods:** This single-center, cross-sectional study involves 100 adult Japanese males with HIV who have received ART for at least 12 months. EA and EAA will be calculated using whole-blood DNA methylation data generated using the Illumina Infinium MethylationEPIC v2.0 BeadChip array and epigenetic clocks developed on Japanese populations. Linear regression models will be used to evaluate the associations between EAA and the factors collected from medical records, laboratory tests, self-administered questionnaires, and comorbidities.

**Discussion:** This study provides baseline data for future longitudinal and comparative studies. The integration of clinical, questionnaire, and omics data facilitates the exploration of biological mechanisms underlying aging in this population. The limitations include the cross-sectional design, which precludes causal inference and the lack of a control group.

**Graphical abstract:** 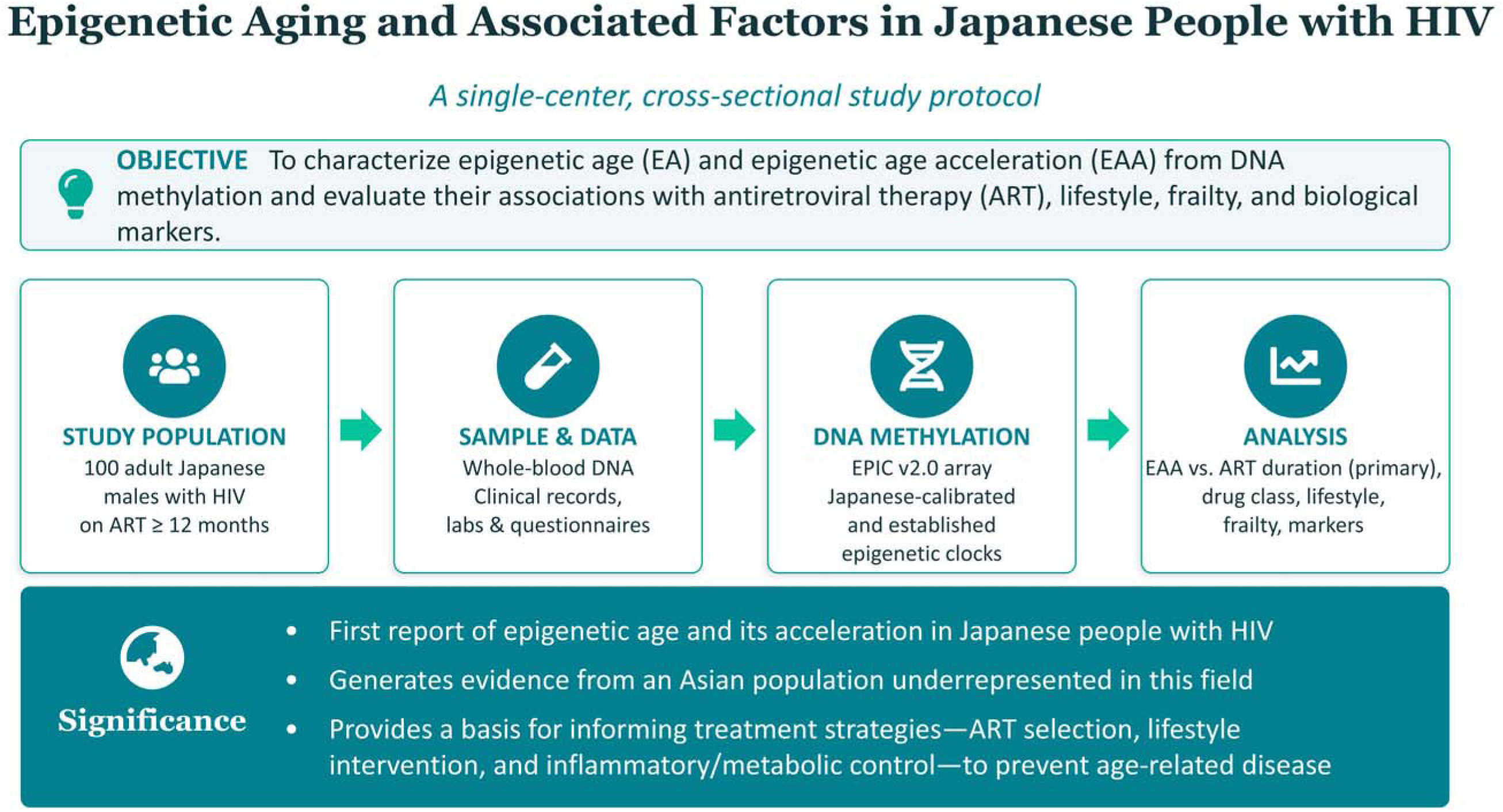

## 1 Background

Human immunodeficiency virus (HIV) infection was once a fatal disease with a poor prognosis following the onset of acquired immunodeficiency syndrome (AIDS). However, rapid advances in antiretroviral therapy (ART), particularly the widespread use of combination therapy, have enabled the long-term suppression of viral load, transforming HIV infection into a manageable chronic condition. Currently, with the early initiation of ART and continuous medication management, the life expectancy of people with HIV (PWH) is approaching that of the general population, and the population of PWH is increasingly aging. In this context, higher incidence rates of age-related non-AIDS-defining illnesses, such as cancer, cardiovascular disease, osteoporosis, and renal dysfunction, have been reported in PWH than in the general population, suggesting that HIV infection is associated with accelerated aging^1^. Aging progression is influenced by multiple factors, including frailty, lifestyle, and immunological and metabolic status^2, 3, 4^.

Epigenetic age (EA), based on DNA methylation, has attracted attention as a biomarker for quantitatively evaluating the progression of biological aging. EA is calculated using statistical models (epigenetic clocks; ECs) that predict age from the methylation proportions of multiple CpG sites. Various ECs have been developed, differing in the definition of age and population used for model training and structure. Specifically, Horvath’s clock and Hannum’s clock—which are trained to predict chronological age—are first-generation epigenetic clocks^5, 6^. PhenoAge, GrimAge, and GrimAge version 2 (GrimAge v2) are second-generation ECs. These clocks were developed to predict year-scale metrics of mortality and morbidity risk by incorporating clinical and molecular biomarkers beyond chronological age^7, 8, 9^. DunedinPACE is known as a third-generation clock and evaluates the pace of aging^10^. Furthermore, population-specific ECs have been developed to account for differences in genetic background, including those for the Japanese population^11^. Epigenetic age acceleration (EAA) is defined as the difference between EA and chronological age or as the residual from regressing EA on chronological age. EA and EAA are used for evaluating anti-aging interventions and exposure, as well as predictors of various health outcomes, including cancer, cardiovascular disease, frailty, and mortality risk^2, 3, 12, 13^.

Previous studies on PWH have reported that EA tends to exceed chronological age, indicating positive EAA^14, 15^. A longitudinal study demonstrated that EAA accumulated during untreated HIV infection, and that this acceleration was greater with longer periods without treatment^15^. However, it has also been observed that EAA may decrease following ART initiation and may become negative in the long term, suggesting that the effects of ART extend beyond viral suppression to the progression of biological aging itself^15^. Furthermore, some studies have suggested that the effects on biological aging may differ between ART drug classes, including associations between specific antiretroviral agents and telomere-related indices or immune exhaustion markers^4, 16^. Nevertheless, most of these findings are based on Western populations, and there is limited evidence regarding EA and EAA in Japanese PWH.

This study aims to evaluate EA and EAA based on DNA methylation data in Japanese PWH and to assess their associations with ART history, frailty, lifestyle factors (smoking, alcohol consumption, exercise, sleep, and dietary habits), and immunological, metabolic, and inflammatory markers using a cross-sectional design. Specifically, the primary objectives of this study are to evaluate EA and EAA in Japanese PWH, and the association between the duration since ART initiation and EAA. Secondary objectives included evaluating the association between EAA and cumulative duration of use for each ART drug class, assessing the associations with frailty and lifestyle factors, and exploring the associations with immunological, metabolic, and inflammatory markers. As this is a single-center cross-sectional observational study without a control group, the focus is on evaluating associations rather than estimating causal effects.

## 2 Methods

### 2.1 Study Design

This is a single-center cross-sectional observational study conducted through a multi-institutional collaboration, enrolling adult Japanese male outpatients with HIV. The study is coordinated by the National Institute of Infectious Diseases of the Japan Institute for Health Security (Tohoku, Tokyo, Japan). The Department of Infectious Diseases at Tokyo Metropolitan Cancer and Infectious Diseases Center, Komagome Hospital (Bunkyo-ku, Tokyo, Japan) is responsible for participant enrollment, collection of clinical information, physical measurements, blood sampling, and collection of self-administered questionnaires. DNA extraction will be performed at SRL, Inc. (Minato-ku, Tokyo, Japan), and DNA methylation analysis will be conducted at Rhelixa, Inc. (Chuo-ku, Tokyo, Japan). Statistical analyses will be performed at the National Institute of Infectious Diseases and the Research Institute of Tuberculosis, Japan Anti-Tuberculosis Association (Kiyose-shi, Tokyo, Japan).

The study period is from the date of institutional approval (November 11, 2025) until March 31, 2030, with the enrollment period scheduled from January 1, 2026, to March 31, 2027.

### 2.2 Participants

To evaluate the status of EA and EAA under homogeneous conditions, this study focused on adult males, who constitute the majority of PWH in Japan. To focus on the stabilized phase of EAA following ART initiation, individuals who received ART for at least 12 months will be included. To avoid conditions that may substantially affect DNA methylation profiles or acute systemic health status, individuals with diseases severely affecting immune function, recent surgery or hospitalization, or viral hepatitis, defined as positive hepatitis B surface antigen (HBsAg) and/or anti-hepatitis C virus (HCV) antibody will be excluded. The inclusion and exclusion criteria are as follows:

#### Inclusion Criteria

1. Diagnosed with HIV infection and currently receiving outpatient care.
2. At least 12 months since ART initiation.
3. Provision of written informed consent.
4. Japanese males aged 18 years or older at the time of providing informed consent.

#### Exclusion Criteria

1. Inability to provide informed consent, complete questionnaires, or provide clinical information, as determined by the attending physician.
2. Diseases with a severe impact on immune function (e.g., active malignancy, autoimmune disease, active tuberculosis, and hematopoietic disorders).
3. History of surgery or hospitalization within the past 3 months.
4. Visit at enrollment for purposes other than routine outpatient care.
5. Positive for HBsAg or anti-HCV.
6. Severe substance abuse or dependence.

### 2.3 Participant Enrollment

At the outpatient clinic, the attending physician will invite patients who potentially meet the eligibility criteria to participate and provide written explanations. Participation in the study is voluntary. After eligibility is confirmed, a participant identification (ID) code will be assigned. A correspondence table linking the ID code to personal information will be strictly managed at the Tokyo Metropolitan Komagome Hospital and will not be shared with other institutions.

The study participation period consists of two visits (visits 1 and 2). At visit 1, the investigators will explain the study, obtain consent, and distribute the self-administered questionnaires. At visit 2, the investigators will collect the questionnaires, perform physical measurements and clinical assessments, conduct medical interviews, and collect blood samples. Information from the medical records, including ART history and historical laboratory data, will be extracted separately. Participants are requested to complete the questionnaire within approximately 7 days prior to visit 2.

### 2.4 Sample Collection and Handling

In addition to clinical blood sampling, whole blood, plasma, and serum samples will be collected for research purposes simultaneously to avoid additional venipuncture.

Whole blood samples collected in EDTA-2Na tubes will be labeled with the participant ID codes and sent to SRL, Inc. for DNA extraction. The extracted DNA will be labeled with the ID codes and transported to Rhelixa, Inc. for methylation analysis. Plasma and serum samples will be aliquoted, labeled with the participant ID code, and transported to the AIDS Research Center at the National Institute of Infectious Diseases, where they will be stored under temperature-controlled conditions.

### 2.5 Information to be Collected

- **Information collected at visit 2**: Chronological age; height, weight, BMI, and clinical frailty scale score; comorbidities; current medications; history of mycobacterial infection and syphilis; ART adherence; smoking history; and alcohol consumption.
- **Clinical laboratory tests at visit 2**: complete blood count with differential, serum biochemistry (e.g., liver and renal function tests, lipid profile, glucose, and HbA1c), inflammatory and coagulation markers (CRP, D-dimer, PT-INR, and APTT), plasma HIV-1 RNA viral load, CD4^+^ T-cell count, CD8^+^ T-cell count, CD4/CD8 ratio, syphilis serology (RPR and TPLA), and QuantiFERON-TB Gold Plus.
- **Information extracted from medical records**: ART initiation date; ART regimen history; history of AIDS-defining illnesses; and detailed clinical information on previous tuberculosis, latent tuberculosis infection, nontuberculous mycobacterial (NTM) infection, and syphilis, including disease type, stage, treatment history, and outcomes. NTM infection is defined as any clinically diagnosed NTM infection with documented follow-up regardless of the site of infection or disease phenotype.
- **Historical laboratory data from medical records**: nadir CD4^+^ T cell count, CD4^+^ T cell count, HIV-1 RNA viral load at diagnosis and peak viral load, mycobacterial culture results, and syphilis serological test results.
- **DNA methylation data**: β-values obtained by the Illumina Infinium MethylationEPIC v2.0 BeadChip array.
- **Tests for research purposes**: immunological, inflammatory, endocrine, metabolic, and other biomarkers (e.g., IL-6, TNF-α, 25(OH)D, soluble PD-L1, and fatty acid profiles) measured using stored plasma and serum samples.
- **Self-administered questionnaires**: Physical activity (International Physical Activity Questionnaire), sleep (Pittsburgh Sleep Quality Index), and dietary habits (Brief-type Self-administered Diet History Questionnaire).

### 2.6 DNA Methylation Analysis

At Rhelixa Inc., genomic DNA derived from whole blood will undergo bisulfite treatment, and DNA methylation will be measured using the Illumina Infinium MethylationEPIC v2.0 BeadChip array (EPICv2). This array contains probes for approximately 930,000 CpG sites. Following measurement, quality control will be performed under standard preprocessing protocols to calculate the methylation proportion (β-value) for each CpG site. This study does not involve whole-genome sequencing.

### 2.7 Calculation of EA

EA will be calculated using multiple ECs. Primary ECs will include the principal component (PC)-based clocks PC-Horvath, PC-Hannum, and PC-PhenoAge, which were developed on the Japanese population^11^. Furthermore, the EA will be calculated using other representative ECs such as the Horvath clock, Hannum clock, PhenoAge, GrimAge, and GrimAge v2. The EAA for each clock will be defined as the residual from the regression of EA on the chronological age. The EAA based on the difference from chronological age will be used for sensitivity analyses. The pace of aging will also be calculated using DunedinPACE software.

### 2.8 Outcome Measures

#### Primary Outcome Measure

The primary outcome measure is EAA calculated based on ECs, particularly those developed for the Japanese population. The primary analysis will evaluate the association between the duration since ART initiation and EAA.

#### Secondary Outcome Measures

Secondary outcome measures include:

1. Association between cumulative exposure to each ART drug class and EAA.
2. Association between EAA and lifestyle factors, physical indicators, frailty, and medical history including comorbidities and infectious diseases.
3. Association between EAA and immunological markers, metabolic indicators, and inflammatory markers.
4. Distribution of EA and consistency among EA values calculated by different ECs.

### 2.9 Sample Size

Assuming a correlation coefficient of −0.30 between the duration of ART and EAA based on previous international studies, 91 cases are required to detect a negative correlation with a 5% significance level and 90% power. The target sample size was set at 100 cases to account for exclusions due to DNA methylation quality control and dropouts due to missing questionnaires or clinical information.

### 2.10 Statistical Analysis

#### Analysis Sets

All participants meeting the eligibility criteria will be defined as eligible. Among the eligible cases, those for which EA or EAA and primary covariates required for the primary analysis are available will be defined as the Full Analysis Set (FAS). Participants with available measurement items for secondary or exploratory analyses comprise the exploratory analysis set. The primary analysis will be conducted on the FAS, and secondary analyses will be conducted on the exploratory analysis set.

#### Statistical Methods

Participant characteristics will be described using means and standard deviations or medians and interquartile ranges for continuous variables and frequencies and percentages for categorical variables. The distributions of EA and EAA will be summarized for each clock and visualized using histograms, scatter plots, or box plots.

For the primary analysis, linear regression models will be used, with EAA as the target variable and duration since ART initiation as the primary explanatory variable. In addition to univariate analysis, multivariate analysis will incorporate clinically important confounders as covariates. Candidate covariates include chronological age, BMI, smoking, alcohol consumption, medical history/comorbidities, HIV-related indicators (e.g., CD4^+^ count, CD4/CD8 ratio, and HIV-RNA viral load), inflammatory markers, and other variables. No additional adjustment for chronological age will be performed when using residual-based EAAs. Analyses will be performed separately for each clock cycle and the estimated regression coefficients with 95% confidence intervals will be presented.

For secondary analyses, the association between the cumulative duration of use of each ART drug class and EAA will be evaluated using linear regression models. As cumulative exposure by drug class may be interdependent and cause statistical issues, models that treat each drug class individually may be considered. Where necessary, analyses will involve categorization based on current regimens, combinations of major drug classes, or the use of regularization methods. The associations between EAA and lifestyle factors, frailty, physical indicators, immune/metabolic/inflammatory markers, and comorbidities, such as syphilis or tuberculosis, will be similarly evaluated. Interaction terms for the duration since ART initiation or major ART drug classes will be introduced to explore the heterogeneity of the associations.

#### Handling of Missing Values

Missing clinical and questionnaire data will be checked based on missing patterns and the application of multiple imputations will be considered if necessary. While the primary analysis will be based on a complete case analysis, sensitivity analyses using multiple imputations will be performed.

#### Adjustment of Multiplicity

The significance level is set at 5% (two-sided). For analyses involving multiple epigenetic clocks and secondary/exploratory analyses, the results will be interpreted using point estimates and 95% confidence intervals. Multiplicity may be addressed through methods for multiplicity adjustment, such as false discovery rate control, if necessary.

#### Sensitivity Analysis

Sensitivity analyses will be performed as necessary, including analyses using models with different sets of covariates or nonlinear models, different approaches to missing data, and comparisons between the difference-based and residual-based EAA definitions.

#### Subgroup Analysis

To examine the factors associated with EAA in detail, subgroup analyses will be conducted based on age, duration since ART initiation, medical history, and lifestyle factors. These analyses will explore the heterogeneity of EAA trends across specific groups. Definitions of the subgroups and statistical analysis methods will be determined based on data distribution and clinical relevance.

#### Exploratory Analysis to Investigate Biological Mechanisms

We will detect differentially methylated positions (DMPs) and differentially methylated regions (DMRs), followed by Gene Ontology analysis of genes proximal to these DMPs and DMRs, to investigate the biological mechanisms.

#### Interim Analysis

No interim analysis is planned in this study.

#### Software

All statistical analyses will be performed using Python (version 3.10 or later) and R (version 4.5 or later).

### 2.11 Ethical Considerations

This study will be conducted in accordance with the Declaration of Helsinki and the Ethical Guidelines for Medical and Biological Research Involving Human Subjects (Japanese National Guidelines). This study was approved by the Institutional Review Board of the Japan Institute for Health Security (Approval No.: JIHS-S-005164-00), and permission was obtained from the head of each participating institution. Participants will be informed about the study’s purpose, methods, voluntary nature of participation, right to withdraw consent, handling of personal information, and expected benefits and risks. Written informed consent will be obtained.

In this study, blood sampling for research purposes will be performed in addition to routine sampling. However, because no new puncture is required, the physical burden is considered minimal. Risks associated with blood collection, such as pain, subcutaneous hemorrhage, or transient discomfort, are comparable to those associated with routine clinical sampling. Personal information will be protected through pseudonymization using participant ID codes, and the correspondence table will be strictly managed at the enrolling institution.

As EA and EAA are currently not established indicators for individual diagnosis or treatment decisions, individual results will not be returned to the participants. The results will be published in academic journals and presented at conferences in a manner such that individuals cannot be identified.

## 3 Discussion

This study evaluates the status of EA and EAA among Japanese PWH and their associations with ART; lifestyle factors; frailty; and immune, metabolic, and inflammatory markers. Beyond its descriptive aims, this study has broader implications for the clinical management of PWH. As different classes of antiretroviral drugs may have differential effects on epigenetic aging^4^ and EAA have been associated with age-related comorbidities and mortality in PWH^17^, the findings of this study could contribute to determining whether biological aging is an important consideration for ART strategies in the future^17^. Furthermore, chronic inflammation is a key driver of both epigenetic aging and age-related diseases, and recent clinical trials have demonstrated that anti-inflammatory therapies can reduce the incidence of such diseases^18^. Given that PWH experience persistent inflammation even under suppressive ART, identifying the determinants of epigenetic aging could inform strategies that combine optimized ART selection, pharmacological anti-inflammatory interventions, and lifestyle modifications. Notably, few studies have evaluated the association between dietary habits and epigenetic aging in PWH, and the detailed dietary assessment in this study addresses this gap. As lifestyle approaches are low-cost and implementable in resource-limited settings, these findings may be relevant to PWH worldwide.

The limitations of this study include its single-center cross-sectional observational design and the absence of a control group, which limits the estimation of causal effects and direct evaluation of temporal changes. Furthermore, because the study population is limited to adult Japanese males, the generalizability of the results to women or other populations is limited. Despite these limitations, this study provides fundamental information that can serve as a basis for future longitudinal and comparative studies.

## Data Availability

The individual-level data generated during this study will not be publicly available because the participants did not provide consent for public data sharing.

## List of Abbreviations

Acquired immunodeficiency syndrome: AIDS
antiretroviral therapy: ART
differentially methylated positions: DMPs
differentially methylated regions: DMRs
epigenetic age acceleration: EAA
epigenetic age: EA
epigenetic clocks: ECs
Full Analysis Set: FAS
hepatitis B surface antigen: HBsAg
hepatitis C virus: HCV
Human immunodeficiency virus: HIV
identification: ID
nontuberculous mycobacterial: NTM
PC: principal component
people with HIV: PWH.

## Study Status

This manuscript is based on the Study Protocol version 1.2 (last updated on December 15, 2025). The first participant was enrolled in the study on March 31, 2026.

## Competing Interests

YT served as a technical advisor in statistical science for Rhelix Inc., from April 2021 to March 2024. YT received a research grant from the Pfizer Health Research Foundation. RN is the founder and chief executive officer of Rhelixa, Inc.

## Funding

This work was supported by the Japan Society for the Promotion of Science (JSPS) KAKENHI (Grant Number: 24K20250), Japan Medical Women’s Association, Pfizer Health Research Foundation, and Research Institute of Tuberculosis.

## Author Contributions

YT, SJ, YKN, and AK conceptualized the study. All authors contributed to study planning. YT and AK developed study protocols. YT and AK developed the statistical analysis plan. YT, YKN, and AK acquired funding. AK was responsible for project administration and supervision. YT and AK drafted the manuscript. All authors reviewed and edited the manuscript and confirmed and approved the final version.

## Notes

### Author Declarations

Ethics committee/IRB of Japan Institute for Health Security gave ethical approval for this work.

